# Repurposing cardiovascular disease risk models to predict incident and co-occurring cardiovascular, cardiometabolic and neurocognitive outcomes

**DOI:** 10.64898/2026.06.07.26355105

**Authors:** Sam Quill, Nish Chaturvedi, Marion van Vugt, Aroon D. Hingorani, Amand F. Schmidt

**Affiliations:** Institute of Cardiovascular Science, University College London, London, UK; British Heart Foundation Centre of Research Excellence, University College London, London, UK; National Institute for Health Research University College London Hospitals Biomedical Research Centre, UK; Department of Cardiology, Amsterdam University Medical Center, Amsterdam, Netherlands

**Keywords:** Risk prediction, cardiovascular disease, cardiometabolic disease, dementia, disease prevention

## Abstract

**Background:** Cardiovascular disease (CVD), cardiometabolic and neurocognitive conditions share risk factors and frequently co-occur. We evaluated whether four established CVD risk prediction models (QRISK3, PCE, SCORE2, SCORE2-OP) can be repurposed to predict 10-year risk of these conditions and their co-occurrence with CVD.

**Methods:** The models were recalibrated using 20% of the UK Biobank (UKB) and evaluated in the remaining 80%. We performed external validation using data from Clinical Practice Research Datalink (CPRD) Aurum, assessing model discrimination (c-statistics) and calibration (intercept and slope). We used permuted feature importance to determine the influence of each individual predictor in the models.

**Results:** Depending on the model, the c-statistics for incident CVD ranged from 0.71 to 0.74 in the UKB test set (16,137 events). Discrimination was equal to or higher than CVD when evaluated against non-traditional CVD outcomes: 0.74 to 0.77 for heart failure (3,471 events), 0.72 to 0.73 for atrial fibrillation (9,213 events), 0.73 to 0.75 for peripheral arterial disease (1,927 events) and 0.80 to 0.82 for abdominal aortic aneurysm (595 events). For the multimorbidity endpoints, model discrimination ranged from 0.74 for the composite of CVD and T2DM (SCORE2-OP) to 0.83 for the composite of CVD and dementia or Parkinson’s disease (QRISK3). When considering the onset of any cardiovascular, cardiometabolic, or neurocognitive outcome discrimination ranged from 0.71 to 0.72. The repurposed models slightly underestimated the predicted risk in the CPRD compared to the UKB: average difference in calibration intercept was at most -0.63. After age and sex, smoking status and systolic blood pressure contributed most to model predictions.

**Conclusions:** Repurposed CVD models can be used to identify 10-year risk of many CVD-related conditions and their multimorbidity. These may be used to support risk-based approaches to prevention and screening. The repurposed models have been made available at: https://repurposed-cvd-risk-models.shinyapps.io/cvd_cmd_dementia_app/

## Background

The global burden of cardiovascular disease (CVD) is expected to rise further over the coming decades, driven by ageing populations, improved survival after index CVD events, and increased prevalence of obesity and type 2 diabetes (T2DM)^1^. As a consequence, the phenotype of CVD increasingly extends beyond myocardial infarction and stroke to include heart failure (HF) and atrial fibrillation (AF)^2^. Due to a combination of shared risk factors and CVD itself causing non-cardiac organ dysfunction, CVD also commonly co-occurs with other chronic conditions. These include T2DM, chronic kidney disease (CKD)^3^, and neurocognitive disorders such as Parkinson’s disease^4^ and dementia^5,6^.

Although these conditions are typically incurable, early and targeted interventions can prevent or delay their onset^7^. Up to 45% of dementia cases could be avoided by addressing modifiable risk factors, such as tobacco exposure, hypertension and dyslipidaemia^5^. Obesity accounts for more than half of T2DM cases worldwide^8^ and, together with hypertension and diabetes, is a leading cause of CKD^9^. Prevention of HF^10^ and AF^11^ similarly targets the same risk factors that drive atherosclerotic CVD, such as cholesterol levels, blood glucose and other elements of Life’s Essential 8^12^.

These CVD-related conditions typically have long prodromal phases before symptoms manifest^10,13^, which is a critical window for prevention. Disease-specific risk prediction models exist, for example for CKD^14^, T2DM^15^ and dementia^16^, but they have not been widely adopted into clinical practice^14–16^. Most currently used CVD risk prediction models are limited to ischaemic heart disease or stroke, despite these no longer being the most common first presentations of cardiovascular disease^2,17^.

Existing CVD models capture many risk factors that associate with these cardiometabolic and neurocognitive outcomes. For example, six of the 15 core modifiable risk factors for dementia outlined in the 2024 Lancet’s report on Dementia Prevention Intervention and Care^5^ (smoking, hypertension, obesity, diabetes, depression, high cholesterol) are included as predictor variables in QRISK3^18^. Given this overlap, we sought to repurpose CVD risk models that are widely used in clinical practice to predict both the incidence of cardiovascular, cardiometabolic and neurocognitive outcomes and their co-occurrence with CVD.

We evaluated the QRISK3, Pooled Cohort Equations (PCE)^19^ and SCORE2/SCORE2-OP^20,21^ risk models on these outcomes using data collected on approximately 500,000 UK Biobank (UKB) participants and over 4.8 million people from Clinical Practice Research Datalink (CPRD) Aurum. The recalibrated, repurposed models have been made publicly accessible: https://repurposed-cvd-risk-models.shinyapps.io/cvd_cmd_dementia_app/.

## Methods

### Data resources

We carried out our primary analysis using data from UKB and externally validated our findings using data from CPRD Aurum. Both data resources have been described in detail elsewhere^22,23^.

The UKB is a longitudinal cohort study that recruited over 500,000 individuals across the UK between 2006 and 2010. These data were randomly split into 20% training and 80% testing sets for model recalibration and evaluation, respectively. To align the external validation cohort with people typically eligible for CVD risk assessment, we limited the CPRD data to participants aged 40 to 84 years at our study start date (1^st^ January 2011).

Linked Hospital Episode Statistics (HES), and Office for National Statistics (ONS) death registry data were available for both UKB and CPRD. For the UKB analysis, we also used data collected at baseline assessment, including demographic characteristics, lifestyle factors, information from nurse-led interviews, and clinical measurements (blood pressure, BMI, and biochemistry data).

### Outcome definitions

CVD was defined as the composite of coronary heart disease (including sudden cardiac death) and ischaemic stroke. We evaluated the broader cardiovascular outcomes of HF, AF, abdominal aortic aneurysm (AAA) and peripheral arterial disease (PAD) individually and as part of an extended composite endpoint, CVD+, consisting of CVD, HF, AF, AAA or PAD.

The individual cardiometabolic outcomes we considered were CKD and T2DM. The neurocognitive outcomes were Alzheimer’s disease, vascular dementia, frontotemporal dementia, Parkinson’s disease, and any type of dementia or Parkinson’s disease.

Additionally, we evaluated CVD-multimorbidity endpoints, where people had a diagnosis of CVD as well as a diagnosis of cardiometabolic disease (CVD and CKD, or CVD and T2DM) or neurocognitive disease (CVD and dementia or Parkinson’s disease).

Finally, we constructed composite endpoint of any major adverse cardiometabolic or neurocognitive outcome (MACNO), defined as the first occurrence of any of the following: CVD+, T2DM, CKD, dementia or Parkinson’s disease. We used any accidental injury (excluding falls) that resulted in an encounter with primary or secondary healthcare services as a negative control. See eTable1 for the code lists used for the outcome definitions.

### Statistical analysis

We applied the original model coefficients from QRISK3, PCE, SCORE2, and SCORE2-OP to calculate predicted risk for each of the models; see eTable2-3 for definitions of the predictor variables. Model performance was assessed using the c-statistic as a measure of model discrimination, and calibration was evaluated using the calibration intercept and slope. All measures are reported with their 95% confidence intervals (CIs).

Dividing the UKB cohort into a 20% training set and an 80% testing set enabled us to recalibrate and independently evaluate each model. We recalibrated the models specifically for each outcome using a rank-preserving transformation, which updated the intercept and slope without influencing model discrimination.

Permuted feature importance analysis was conducted by randomly shuffling each feature 50 times and recording the average change in c-statistic(16).

### Sensitivity and subgroup analyses

For the individual neurocognitive outcomes, we performed additional analyses to investigate whether the model predictions were robust to individuals with different levels of genetic risk. We stratified participants by *APOE4* carrier status in the UKB cohort using whole genome sequencing data. We compared three types of carriership: homozygous ε4 carriers (ε4/ε4), heterozygous ε4 carriers (ε3/ε4 or ε2/ε4), and those carrying at least one ε4 allele (combining homozygous and heterozygous carriers).

For the multimorbidity endpoints, we investigated whether the temporal order in which individual conditions developed influenced model performance, for example, CVD after CKD compared to CKD after CVD.

The primary analysis was limited to participants with complete data; see eTable4 for details of missing data on model variables in UKB and CPRD. We repeated our analyses after imputing missing data in the CPRD cohort using the *missRanger* R package(13).

## Results

### Descriptive results

Data were available on 502,126 participants from UKB and 4,810,089 participants from CPRD. The mean age was 56.6 years (standard deviation (SD): 8.1) in UKB and 58.5 (SD: 12.1) in CPRD; 273,158 (54.4%) and 2,404,320 (50%) were female, respectively. The median follow-up for both UKB and CPRD was 10 years (UKB quartile(Q)1;Q3: 10; 10, CPRD Q1;Q3: 6.1; 10); Table 1.

**Table 1.** Baseline characteristics of participants in UK Biobank and CPRD cohorts. *Baseline characteristics are presented as mean (standard deviation) for continuous variables and frequency (percentage) for categorical variables. The percentage of missing data is shown for each variable within each cohort. BMI, body mass index; HDL cholesterol, high-density lipoprotein cholesterol; UKB, UK Biobank; CKD, chronic kidney disease; CPRD, Clinical Practice Research Datalink; Q1;Q3, quartile 1; quartile 3*.

| Variable | UKB Mean (SD) or N (%) | UKB Missing (%) | CPRD Mean (SD) or N (%) | CPRD Missing (%) |
| --- | --- | --- | --- | --- |
| Overall sample size | 502,126 |  | 4,810,089 |  |
| **Follow-up time in years (median, Q1/Q3)** | 10 (10; 10) |  | 10 (6.1; 10) |  |
| **Age (years)** | 56.6 (8.1) | 0 (0%) | 58.5 (12.1) | 0 (0%) |
| **Female sex (UKB), gender (CPRD)** | 273,158 (54.4%) | 0 (0%) | 2,404,320 (50%) | 24 (<0.1%) |
| **Mean systolic blood pressure (mmHg)** | 138 (18.7) | 1,318 (0.3%) | 132 (15.7) | 760,049 (15.8%) |
| **Total cholesterol (mmol/L)** | 5.7 (1.1) | 32,874 (6.5%) | 5.1 (1.1) | 1,744,818 (36.3%) |
| **HDL cholesterol (mmol/L)** | 1.4 (0.4) | 72,567 (14.5%) | 1.4 (0.4) | 2,037,512 (42.4%) |
| **BMI (kg/m<sup>2</sup>)** | 27.4 (4.8) | 3,328 (0.7%) | 27.8 (5.6) | 1,264,154 (26.3%) |
| **Personal history of:** |  |  |  |  |
| **CVD** | 33,277 (6.6%) |  | 598,457 (12.4%) |  |
| **Dementia or Parkinson's Disease** | 784 (0.2%) |  | 54,471 (1.1%) |  |
| **Diabetes** | 30,781 (6.1%) |  | 632,208 (13.1%) |  |
| **CKD** | 2,399 (0.5%) |  | 358,863 (7.5%) |  |
| **Ethnicity:** |  | 895 (0.2%) |  | 247,082 (5.1%) |
| **White or not stated** | 474,238 (94.4%) |  | 4,160,043 (86.5%) |  |
| **Indian** | 5,944 (1.2%) |  | 90,799 (1.9%) |  |
| **Pakistani** | 1,833 (0.4%) |  | 39,744 (0.8%) |  |
| **Bangladeshi** | 236 (<0.1%) |  | 14,491 (0.3%) |  |
| **Other Asian** | 2,687 (0.5%) |  | 49,864 (1.0%) |  |
| **Black Caribbean** | 4,509 (0.9%) |  | 36,491 (0.8%) |  |
| **Black African** | 3,390 (0.7%) |  | 68,997 (1.4%) |  |
| **Chinese** | 1,573 (0.3%) |  | 17,562 (0.4%) |  |
| **Other ethnic group** | 6,821 (1.4%) |  | 86,624 (1.8%) |  |
| **Smoking status:** |  | 19,745 (3.9%) |  | 588,137 (12.2%) |
| **Non-smoker** | 273,336 (54.4%) |  | 2,351,207 (48.9%) |  |
| **Ex-smoker** | 172,923 (34.4%) |  | 1,234,591 (25.7%) |  |
| **<10 cigarettes/day** | 7,207 (1.4%) |  | 200,791 (4.2%) |  |
| **10-19 cigarettes/day** | 15,420 (3.1%) |  | 255,872 (5.3%) |  |
| **>19 cigarettes/day** | 13,495 (2.7%) |  | 179,491 (3.7%) |  |

During follow-up 23,660 (5.9%) and 224,799 (6.6%) CVD events occurred in UKB and CPRD, respectively. CVD+ (CVD definition that includes AF, HF, AAA and PAD) occurred in 35,654 (8.9%) people in the UKB and 375,737 (10.4%) in CPRD. In UKB and CPRD, incident T2DM occurred in 15,296 (3.3%) and 227,760 (5.5%), CKD in 23,341 (4.7%) and 354,869 (8.0%), and incident dementia or Parkinson’s Disease occurred in 6,287 (1.3%) and 189,168 (4.0%) respectively. Full counts of incident cases for each condition are provided in eTables5-6.

### Discrimination for individual CVD, cardiometabolic, and neurocognitive outcomes

The c-statistics for CVD at 10-years were 0.74 (95%CI 0.73; 0.74) for QRISK3, 0.72 (95%CI 0.72; 0.73) for PCE and SCORE2, and 0.71 (95%CI 0.71; 0.72) for SCORE2-OP. For the single-outcome endpoints, the c-statistics ranged from 0.58 (95%CI 0.55; 0.60) for PCE predicting subarachnoid haemorrhage to 0.83 (95%CI 0.82; 0.84) for QRISK3 predicting vascular dementia; Figure 1 and eTable7. The discrimination of the CVD models for HF, AF, PAD, and AAA (c-statistics all 0.72 or greater) was comparable to or higher than their original atherosclerotic CVD endpoints. Discrimination was also higher for dementia than for CVD, particularly vascular dementia (median difference 0.09, 95%CI 0.07; 0.10). For T2DM, the model which had discrimination closest to CVD was QRISK3 (c-statistic: 0.71 95%CI 0.70; 0.71). All four models also showed CVD-equivalent discrimination for CKD; c-statistics ranged from 0.71 (95%CI 0.71; 0.71) for the PCE model to 0.74 (95%CI 0.74; 0.75) for QRISK3. The median c-statistic for accidental injury, the negative control outcome, was 0.53 (Q1;Q3 0.53; 0.54).

**Figure 1.**
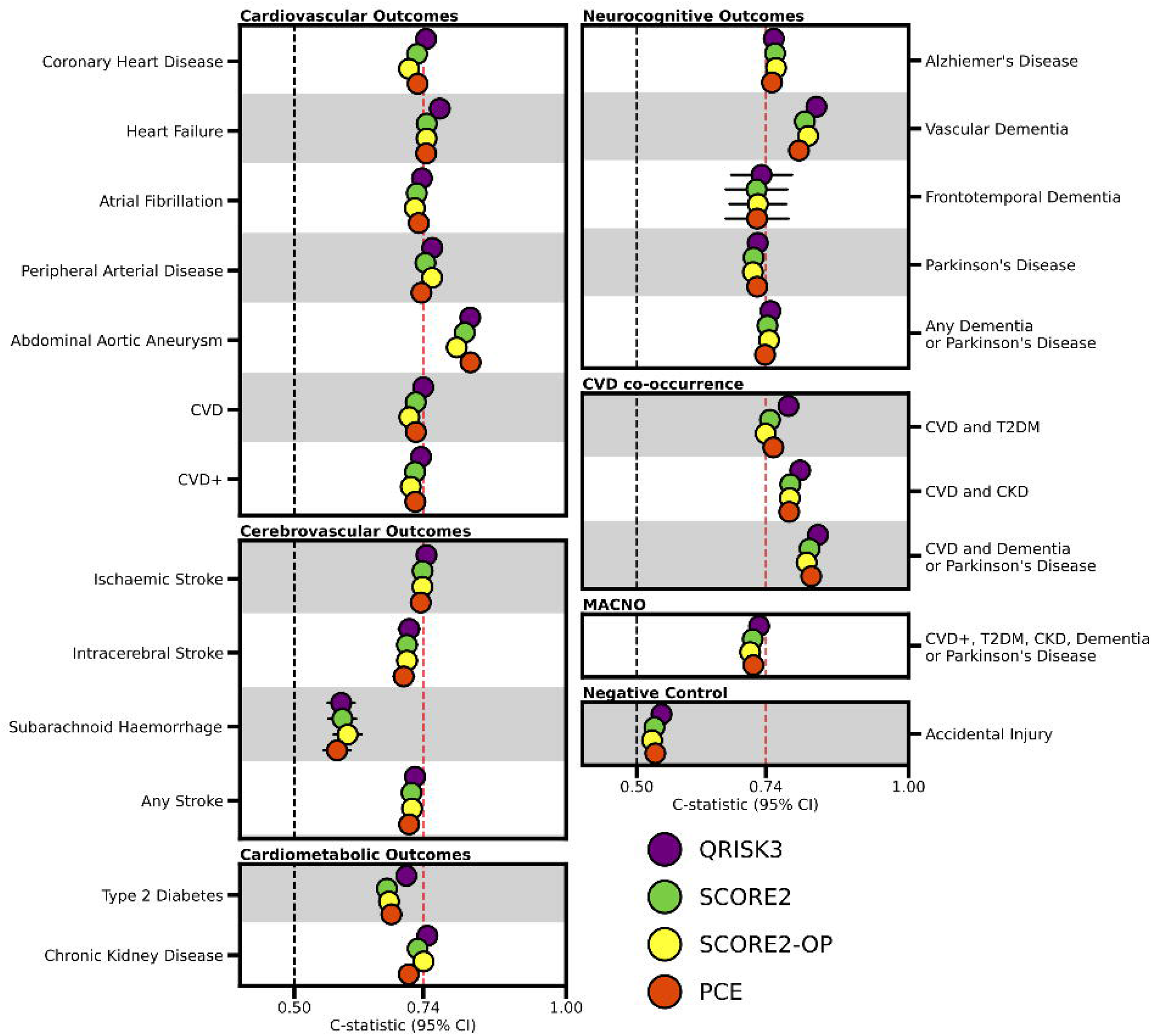
Discrimination of CVD risk prediction models for incident cardiovascular diseases, cardiometabolic diseases, neurocognitive outcomes, CVD-multimorbidity and accidental injury (negative control). *The discrimination of the four repurposed CVD models for the CVD, cardiometabolic, neurocognitive, composite and multimorbidity outcomes using data from the UK Biobank. Accidental injury is included as a negative control. The red, vertical dashed line corresponds to the performance of QRISK3 for CVD at 10 years (c-statistic of 0*·*74, 95%CI 0.73; 0.74). The vertical dashed black line indicates the c-statistic value of 0*·*5. For each outcome, people with diagnoses related to the outcome being predicted at the study (e.g. pre-existing CKD when predicting incident CKD) were excluded (eTables5-6). Abbreviations: 95% CI, 95% confidence interval; CKD, chronic kidney disease; CVD, cardiovascular disease; MACNO; major adverse cardiometabolic or neurocognitive outcome; T2DM, type 2 diabetes mellitus*.

### Discrimination for CVD-multimorbidity and composite outcomes

The c-statistics for the CVD-multimorbidity endpoints ranged from 0.74 (95%CI 0.72; 0.75) for CVD and T2DM using SCORE2-OP to 0.83 (95%CI 0.82; 0.85) for CVD and dementia or Parkinson’s disease using the QRISK3 model; Figure S1 and eTable7. Model discrimination for the three multimorbidity outcomes all exceeded the discrimination for CVD itself; Figure 1. The order of index disease had little influence on discrimination; the models performed slightly better when the non-CVD disease preceded CVD (median difference 0.02 95%CI 0.01; 0.03). The discrimination was almost identical for the composite endpoints (CVD+ and MACNO) as for CVD; median difference: 0.01 (95%CI 0.00; 0.01), eTable7.

### Performance of the recalibrated models

After recalibrating the CVD models to the UKB training set, QRISK3 was the best calibrated model for the individual outcomes (median calibration intercept: -0.01 (Q1;Q3: -0.07; -0.03), median calibration slope: 1 (Q1;Q3 0.96; 1.09) and the CVD-multimorbidity outcomes (median calibration intercept: 0.06 (Q1:Q3: 0.05; 0.06), median calibration slope: 1.02 (Q1;Q3: 1.01; 1.08); Figure 2, eTable7. All four CVD models were nearly perfectly calibrated for the CVD+ and MACNO composite endpoints (median calibration intercept: -0.02 (Q1;Q3: -0.02; -0.02), median calibration slope: 1.00 (Q1;Q3: 0.98; 1.03); eTable7).

**Figure 2.**
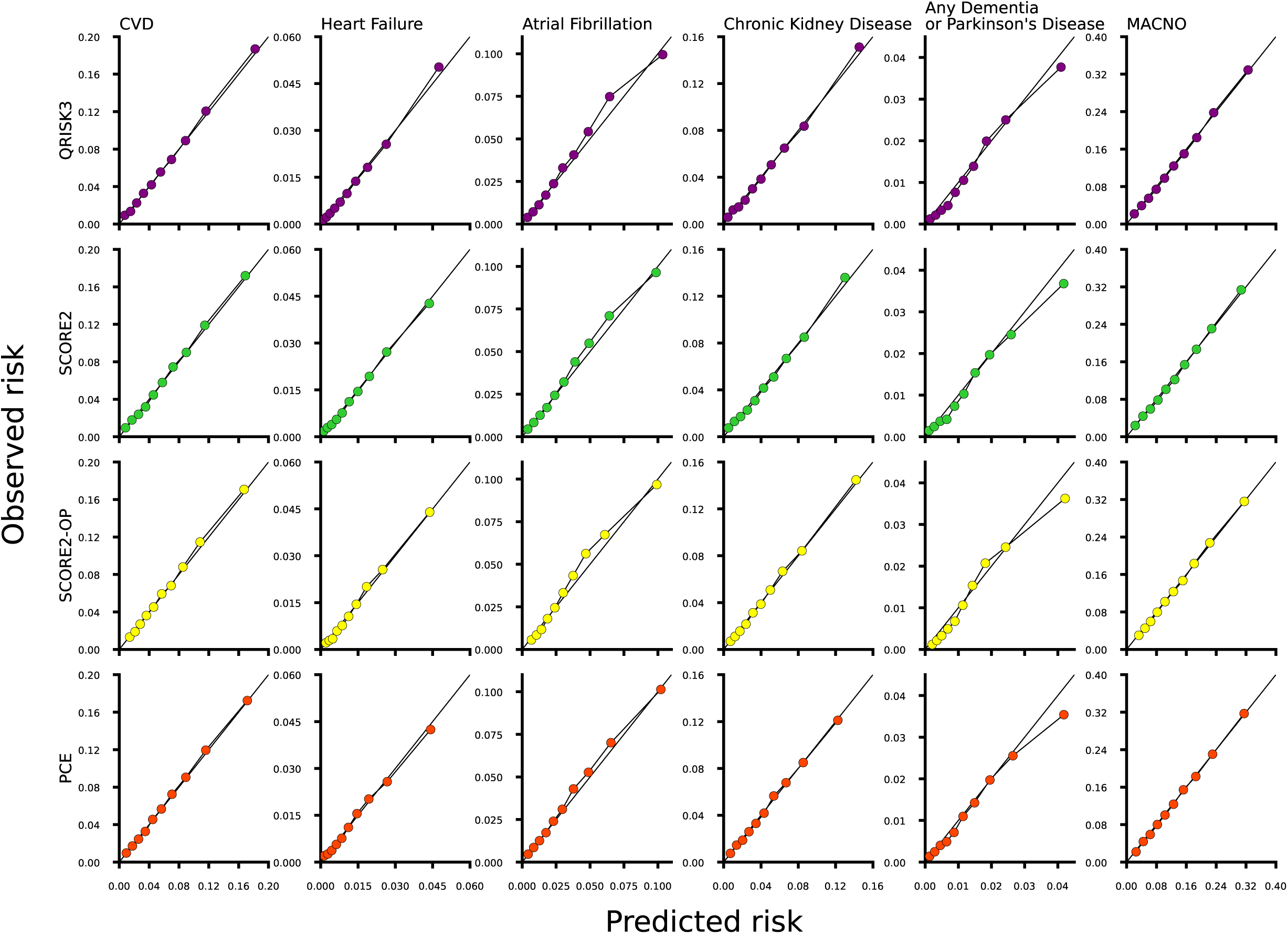
Calibration plots comparing the repurposed CVD risk models for CVD, heart failure, atrial fibrillation, chronic kidney disease, dementia or Parkinson’s disease and any major adverse cardiometabolic or neurocognitive outcome (MACNO) in UK Biobank data after model recalibration. *Agreement between the predicted risks (x-axis) and observed risks (y-axis) of the four CVD risk models (QRISK3, PCE, SCORE2, SCORE2-OP) for six outcomes; CVD, heart failure, atrial fibrillation, chronic kidney disease, any dementia or Parkinson’s disease, and any major adverse cardiometabolic or neurocognitive outcome (MACNO). Each point corresponds to a risk decile and the solid diagonal line to perfect calibration. For each outcome, participants with prevalent conditions were excluded; eTables12. The calibration plots for all outcomes considered in this study can be found in Supplemental Figures S5-9. Abbreviations: MACNO, any major adverse cardiometabolic or neurocognitive outcome*.

### External validation in CPRD

The CVD models showed generally higher discrimination but were less well calibrated in CPRD than in UKB; Figure 3, eTable7. The discrimination for each of the models was highly correlated between UKB and CPRD; Figure 3. Spearman’s correlation (r) ranged from 0.79 for PCE to 0.88 for SCORE2-OP. The model with the highest discrimination was the SCORE2 model when predicting vascular dementia, which had a c-statistic of 0.86 (95%CI 0.85; 0.86). All four CVD models tended to underestimate risk in the CPRD cohort compared to the UKB cohort. The median of the differences between calibration intercepts for each outcome in UKB and CPRD ranged from -0.63 for QRISK3 (p=0.001) to -0.18 for SCORE2-OP (p=0.05), and from -0.35 for SCORE2 (p<0.001) to -0.07 for SCORE2-OP (p=0.05) for the calibration slope; Figure 3. The models were reasonably well calibrated for many outcomes, e.g. for Parkinson’s disease, the PCE model had calibration intercept of 0.13 (95%CI 0.11; 0.14) and calibration slope of 0.96 (95%CI 0.95;0.98); eTable7.

**Figure 3.**
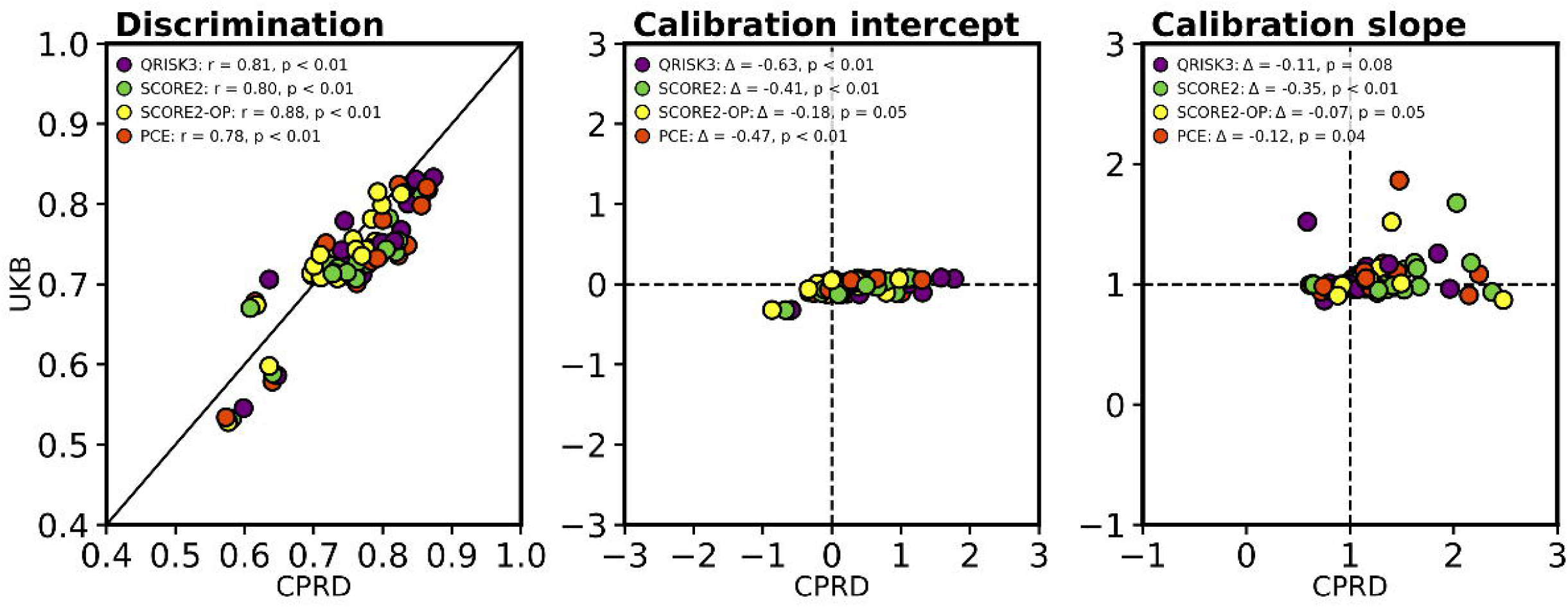
A comparison of the discrimination and calibration of each model between the UK Biobank and CPRD datasets. *The correlation between model performance metrics (discrimination, calibration intercept, and calibration slope) comparing UK Biobank to CPRD. The diagonal line in the discrimination plots (left column) indicates perfect correlation. The intersection of the dashed lines in the calibration intercept and slope plots (middle and right columns) corresponds to perfect agreement across datasets. The colours of the dots indicate each of the models evaluated (see legend). Participants with missing predictor or outcome data were excluded from both cohorts. For each outcome, individuals with prevalent disease were also excluded (see eTables5-6). Abbreviations:* Δ*, median of the differences between estimates in UKB and CPRD (UKB minus CPRD) for each outcome; CPRD, Clinical Practice Research Datalink; UKB, UK Biobank*.

**Figure 4.**
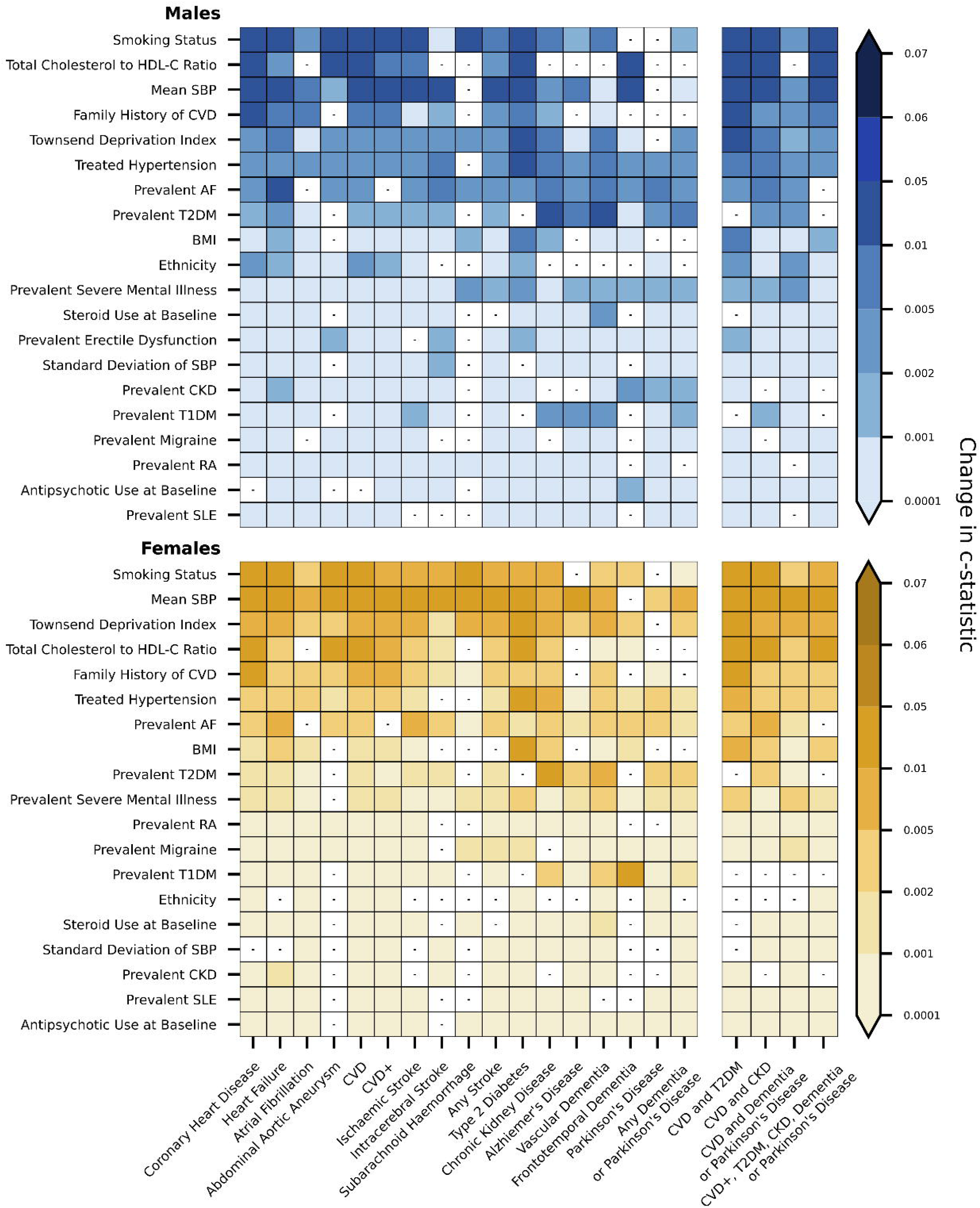
Feature importance in the QRISK3 model for cardiovascular diseases, cardiometabolic diseases, neurocognitive conditions, composite endpoints, and multimorbidity endpoints. *The feature importance results for the QRISK3 model in the UKB for each outcome, measured as the change in c-statistic when each variable is randomly permuted. The top panel (blue) shows results for male participants, and the bottom panel (yellow) shows results for female participants. Predictor variables (y-axis) are ordered by overall importance across all outcomes. The darker shading indicates greater feature importance. The white tiles with a small, black horizontal line indicate which features did not contribute to the c-statistic in the QRISK3 model for that outcome. Abbreviations: AF, atrial fibrillation; BMI, body mass index; SBP, systolic blood pressure; CKD, chronic kidney disease; CVD, cardiovascular disease; HDL-C, high-density lipoprotein cholesterol; RA, rheumatoid arthritis; SLE, systemic lupus erythematosus; T1DM, type 1 diabetes mellitus; T2DM, type 2 diabetes mellitus*.

### Sensitivity analysis

For the neurocognitive outcomes, there was little difference in discrimination by *APOE4* carrier status; eTable8, Supplementary Figure S1. For many of the outcomes we considered, model discrimination was similar in the complete-case and imputed CPRD cohorts. The difference in c-statistic for CVD was 0.002 and overall, the median difference was 0.04 (Q1;Q3: 0.01; 0.09); eTable9.

### Feature importance

For QRISK3, the most influential predictors of CVD were smoking status, mean SBP, and total cholesterol to high-density lipoprotein cholesterol (TC:HDL-C) ratio. For CVD+, mean SBP was most influential in women and both mean SBP and smoking status were in men. For T2DM, the most influential predictors in women were BMI, treated hypertension, TC:HDL-C ratio, Townsend Deprivation Index, and mean SBP; in men, smoking status was more important than BMI. T2DM was the most influential predictor of CKD. For any dementia or Parkinson’s disease, the most influential predictor was mean SBP in women and T2DM in men. For MACNO, mean SBP and TC:HDL-C ratio were the most influential features in both men and women, as was smoking status in men. In the PCE, SCORE2 and SCORE2-OP models, mean SBP was the most important predictor for the majority of conditions; Supplementary Figures S2-4.

## Discussion

We have successfully repurposed clinically used CVD risk models to predict non-traditional cardiovascular, cardiometabolic and neurocognitive outcomes, including CVD-related multimorbidity outcomes. The recalibrated models showed similar discriminative performance and reasonable calibration in the CPRD cohort used for external validation.

All the models we evaluated discriminated well for both the single and composite outcomes. The highest discrimination for an individual (vascular dementia), composite (CVD+), and multimorbidity endpoint (CVD and dementia or Parkinson’s disease) were all achieved by QRISK3. Model calibration varied from poor (QRISK3 for frontotemporal dementia) to excellent (PCE for chronic kidney disease). In general, the composite outcomes were better calibrated than the individual or the multimorbidity endpoints.

For both men and women, we found that the most influential model predictors after age were smoking status, SBP, and serum cholesterol levels (TC:HDL-C ratio in QRISK3; high-density lipoprotein (HDL) cholesterol in PCE, SCORE2 and SCORE2-OP). Model feature importance does not necessarily indicate causal relationships between predictors and outcomes, nor do we intend our results to be interpreted as such. However, the factors used in the models may associate with common causal pathways shared by multiple diseases.

Our findings are relevant to preventing cardiometabolic disease and its complications, including CVD, neurocognitive diseases and multimorbidity. The majority of risk prediction models currently used for primary prevention focus on narrow disease endpoints^24^. To estimate an individual’s risk of multiple conditions, clinicians are required to calculate multiple separate risk scores, which is time-consuming and often impractical in busy clinical environments. Multi-outcome models are not routinely available and those in development have not undergone extensive validation. Previous studies have shown that risk models using data collected during routine health checks can predict various cardiovascular, renal and cognitive endpoints, including HF, CKD and dementia^24,25^. The PCE model has been shown to predict incident HF^26^ and SCORE2 demonstrates good discrimination for all cause dementia^27^. However, none of these studies performed external validation^24–27^ or provided accurate recalibrated prediction rules^27^. None investigated whether the models could predict CVD-multimorbidity.

The AHA’s PREVENT algorithms have expanded their scope of CVD risk prediction to include heart failure^28^, which has led to a renewed focus on heart failure prevention^10^. We demonstrate that this is possible for a much wider set of cardiometabolic, neurocognitive and multimorbidity endpoints without developing new models from scratch or adding new predictors to existing models.

We found that our repurposed CVD models performed comparably or better than purpose-built tools for conditions where risk models are already widely used in clinical practice. For HF, the QRISK3 model showed similar discrimination to the published results of the PREVENT-HF algorithm^28^. For AF, QRISK3 showed higher discrimination than commonly used models such as CHARGE-AF, HARMS_2_-AF and CHA DS -VASc^29^. All four repurposed CVD models performed similarly to existing 10-year risk prediction tools for dementia^16^. Furthermore, the repurposed CVD models demonstrated high discrimination and good calibration on external validation for many conditions where disease-specific prediction tools are not currently used in primary prevention, including CKD, AAA, and PAD.

With a more comprehensive view of overall disease risk, these models could shape how we set thresholds for prevention. For individuals at moderate risk of any one of several related conditions, proactive risk management may be more justified than when considering a single disease in isolation. Similarly, those identified as being at risk of developing several conditions in different organ systems may warrant even more intensive risk factor modification. Given the pleiotropic effects of many available cardiovascular drugs, pharmacological therapy may soon play a larger role in preventing multiple CVD-related diseases. For example, sodium-glucose cotransporter-2 (SGLT2) inhibitors slow CKD progression and reduce CVD risk in people with and without diabetes^31–33^.

Public awareness of the modifiable risk factors for many CVD-related conditions is low, including for CKD and dementia^35,36^. Informing individuals of their CVD risk has been shown to improve uptake and adherence to preventative treatment^37^. By linking well-known cardiovascular risk factors to non-CVD conditions, the repurposed models could help to support communication about the many benefits of risk factor modification for long term health.

As well as supporting primary prevention, the repurposed CVD models could be used in risk-stratified screening for CVD-related disease and multimorbidity. Few systematic screening programmes currently exist for these conditions. When they do, eligibility for screening is often based on relatively crude cutoffs, such as age^38^ or age, sex, and occasionally smoking history^39^. The repurposed CVD models may be used to identify high-risk individuals as candidates for targeted screening, including echocardiography to detect pre-clinical heart failure^28^, ultrasound imaging to screen for AAA^40^, or point-of-care testing for CKD in community settings^41^. This may facilitate screening for conditions where population-wide approaches are currently unfeasible.

Similarly, the repurposed CVD models could complement emerging risk-based screening programmes. For instance, although blood-based biomarkers for predicting incident dementia are not currently indicated for population-level screening, combining these biomarkers with an initial risk-stratification step could improve early identification of individuals at risk of both CVD and dementia^42^.

This study has some limitations. First, our primary analysis evaluated model performance at 10 years of follow-up; longer time frames may be relevant for certain outcomes, such as dementia. However, although midlife is an important time for intervention, modifying risk factors in older adults may still reduce dementia risk^5^. Second, model performance was largely consistent between complete case and imputed analyses, but discrimination was attenuated a small number of conditions; the greatest reductions were seen for the peripheral arterial disease, abdominal aortic aneurysm, and Parkinson’s disease endpoints (difference in c-statistic of 0.22, 0.17, 0.13 respectively). Third, a vast number of CVD risk scores have been published to date and not all were evaluated in this study. However, considering we found minimal differences between the four models we evaluated, and that most CVD risk models use a similar set of predictor variables^43^, we would anticipate finding similar results for other models.

A core strength of this study is that we repurposed CVD models that are already commonly used in existing electronic health record systems. This approach circumvents many of the barriers that typically impede adoption of new risk models into clinical practice^44,45^. The required predictor variables are often routinely collected, and the models are both familiar to clinicians and recommended by health authorities worldwide^7,46,47^. Our external replication in nationally representative data provides evidence of how these models are likely to perform in clinical practice.

## Conclusion

Four CVD risk models available in most primary care settings (QRISK3, PCE, SCORE2, SCORE2-OP) can be repurposed to predict AF, HF, PAD, AAA, T2DM, CKD, dementia, and Parkinson’s disease. These models can furthermore identify individuals at risk of CVD multimorbidity, such as CVD with dementia. Smoking status and systolic blood pressure were the most important predictors in the models, after age and sex. These repurposed CVD models may support risk-based prevention and screening for CVD-related diseases and multimorbidity. The repurposed models are available as a web application at: https://repurposed-cvd-risk-models.shinyapps.io/cvd_cmd_dementia_app/.

## Availability of data and materials

Data are available from the UK Biobank and the Clinical Practice Research Datalink, conditional on successful application.

## Author contributions

SQ, NC, AH and AFS designed the study. SQ performed the analyses and drafted the manuscript. NC, AH and AFS all provided critical input on the analysis, as well as on the drafted manuscript.

## Supporting information

Supplemental eTables

## Acknowledgments

This research has been conducted using the UK Biobank Resource under Application Number 12113. The authors are grateful to UK Biobank participants. This publication is part of the project “Computational medicine for cardiac disease” with file number 2025.027 of the research programme “Computing Time on National Computer Facilities” which is (partly) financed by the Dutch Research Council (NWO). The authors acknowledge the use of the UCL Myriad High Performance Computing Facility (Myriad@UCL), and associated support services, in the completion of this work. The authors would also like to thank Mr. Simon Parker for his work developing code that was used to process and validate CPRD Aurum data in this study.

## Code availability

All analyses were performed using R version 4.3.1. The figures were created using matplotlib, plot-misc 2.2.0^48^, and Python version 3.1.1. The repurposed, recalibrated models have been made available at: https://repurposed-cvd-risk-models.shinyapps.io/cvd_cmd_dementia_app/

