## Supplemental Materials for "Repurposing cardiovascular disease risk models to predict incident and co-occurring cardiovascular, cardiometabolic and neurocognitive outcomes"

### Supplementary Methods:

#### Data resources:

The longitudinal UK Biobank (UKB) cohort study enrolled over 500,00 participants between 37-73 years of age from 22 sites in the UK. Recruitment was conducted between 2006 and 2010. We extracted data collected at participants' first UKB study visit, included demographic, lifestyle, biometric, and clinical data such as blood pressure and cholesterol measurements. We used data from the self-reported questionnaires to define the family history of cardiovascular disease definition. We used this data from the self-reported questionnaire to supplement the EHR-derived definitions of prevalent conditions. We also used self-reported data from the UKB nurse-led interviews that were conducted during the index UKB study visit to define personal medical history, medication use at baseline and prior clinical procedures.

We additionally used data from participants' linked primary and secondary care electronic health records (EHR) to define the predictor variables for the models. Secondary care EHR (Hospital Episode Statistics (HES))) data and from Office of National Statistics death registry records[1] were available for all consenting UKB participants. Data from primary care records were available for approximately half of the UKB participants. Code lists used to define the variables used in this analysis be found in eTables1-3.

The replication cohort was derived using data from Clinical Practice Research Datalink (CPRD) Aurum. CPRD Aurum includes records for all individuals who were registered with participating GP practices using EMIS Web® software. Any individuals registered between 1st April 1997 and 29th March 2021 were potentially eligible for inclusion in our study, corresponding to the availability of linked secondary care (HES) and mortality (ONS death registry) data in CPRD. Individuals without linked data[1] or did not meet CPRD's data quality standards were excluded[2]. We excluded individuals in CPRD who's gender was documented as neither male nor female (n=24, <0.0001%; eTable4) because the CVD models used in this study were not developed to accommodate sex or gender data other than biological sex assigned at birth[3]. To increase likelihood that pre-existing diagnoses were captured in an individual's EHR, we required individuals to be registered with their primary care practice for a minimum of one year prior to the study start date (1st January 2011) in order to be considered eligible for inclusion[4]. For both UKB and CPRD, we defined a one-month clearance period after the study start date to reduce likelihood of prevalent disease being classified as incident[5]. Participants who died or were lost to follow-up during this time (i.e. before the start of the follow-up) were excluded from UKB (n=44). Similarly, individuals in CPRD who deregistered, died, or had no data collected before follow-up began were excluded (n=67,768).

The number of people excluded at each stage of the data processing pipeline for each outcome is summarised in the supplementary tables eTable5, eTable6 and eTable11 for UKB, CPRD, and the imputed CPRD cohort, respectively. Example flowcharts detailing the exclusion counts at each stage when preparing the cohorts for predicting chronic kidney disease are presented in Supplementary Figures S11-12 for UKB and CPRD respectively. The prevalent disease definitions and baseline medications used to identify pre-existing disease are detailed in eTable12. For many outcomes, we defined the set of pre-existing conditions to be excluded more broadly than the specific outcome being predicted. This was to exclude individuals with closely related or potentially misdiagnosed pre-existing conditions. For example, individuals with any dementia diagnosis or taking dementia-related medications at baseline were excluded when predicting incident Alzheimer's dementia. For outcomes where medications were not specific enough for the outcome being predicted, no medications were used to define prevalent conditions, e.g. beta blockers or direct oral anticoagulants were not used as a proxy for prevalent atrial fibrillation. We restricted our analysis to participants with no missing data in any of the models. This to enable the performance of the models to be compared on the same set of participants for each outcome.

The descriptive statistics for the UKB and CPRD cohorts (including the CPRD cohort after imputing missing data) can be found in eTables13-14.

**Predictor variable definitions:**

We defined baseline medication use as at least two prescriptions and the most recent prescription being within 1 month of the start of the study in CPRD. The self-reported medication use survey in UKB did not document the timing of treatment so we assumed that participants who reported relevant medication use at their index UKB visit were currently using these medications. As the age at which the family member experienced CVD was not recorded, we assumed this occurred under 60 years of age. In CPRD we assumed that no documented evidence of family history of CVD indicated no relevant family history of heart disease in first degree relatives under 60 years of age.

In CPRD, the ethnicity variable was first defined using HES data. If this data were missing, we used ethnicity data from the CPRD observation table to supplement it. Similarly in CPRD, we used individual level Index of Multiple Deprivation (IMD), rather than Townsend Deprivation Index. When data on individual-level IMD was not available, the IMD level of the primary care practice the individual was registered with was used instead.

The medcodes we used to define the clinical measurements were created using CPRD's medcode dictionary guided by publicly available lists on the OpenCodelists website[6]. Implausible values were excluded for the clinical measurements in UKB and CPRD; <70 mmHg or >270 mmHg for mean systolic blood pressure; <9 kg/m² or >92 kg/m² for BMI (based on minimum and maximum height and weight values); <1 mmol/L or >20 mmol/L for total cholesterol; <0.1 mmol/L or >5 mmol/L for HDL cholesterol.

**Outcome definitions:**

In the UKB, we used all data collected at the index study visit, as well as data from linked EHR sources to defined prevalent disease. However, since some of these data resources were not repeated throughout follow up (e.g., the nurse-led surveys and questionnaires were only completed at participants' first UKB visit) incident events were defined using the linked primary and secondary care data sources only.

We sourced initial disease code lists from published code lists repositories such as the Health Data Research UK Phenotype Library [7]. SQ reviewed these code lists, supplemented them with additional codes if relevant codes or code types were missing (such as OPCS-4 procedure codes). The ICD-10 codes were mapped to ICD-9 and Read codes using the UKB mapping files[8].

For the replication analysis in CPRD, the code lists used to define conditions in UKB were mapped to SNOMED-CT codes based on mapping files from the NHS Technology Reference Update Distribution (TRUD) website. We then converted the SNOMED-CT to CPRD Aurum medcode IDs using CPRD Aurum's medcode dictionary. Codes that did not map correctly were replenished by adding relevant medcodes found through searching the medcode dictionary manually and manually. The same ICD10 codes and OPSC-4 procedure codes were used in the primary and replication cohorts. SQ reviewed all code lists used in the study prior to the analyses.

We used CVD (the composite of coronary heart disease (including sudden cardiac death) and ischaemic stroke) as a positive control and accidental injury as a negative control in our analyses. We excluded falls from the definition of accidental injury because we assumed falls could be associated with the model inputs or the outcomes being predicted[9]. Compared to our definition of CVD, the four models we evaluated in this study were developed using slightly different definitions of CVD. In addition to coronary heart disease and ischaemic stroke, the QRISK3 model included transient ischaemic attack (TIA) in its definition of CVD[10]. Although none of the other three models were trained against a CVD definition that included TIA, all used any type of stroke instead of ischaemic stroke[11,12].

The composite endpoints we constructed (CVD+, MACNO) were defined as the first occurrence of any of their constituent disease endpoints. In contrast the multimorbidity outcomes (CVD and T2DM, CVD and CKD, CVD and dementia or Parkinson's disease) were defined as the earliest date of the second disease endpoint. Individuals with evidence of any constituent condition in the composite or multimorbidity endpoints at baseline were excluded.

**Statistical analysis:**

The number of participants remaining after applying the exclusion criteria differed according to the endpoint being predicted, e.g. excluding pre-existing disease related to the outcome of interest. We constructed outcome-specific training (20%) and testing (80%) sets using simple random sampling, so that each of the four models were compared on the same participants for a given outcome.

We evaluated the models based on the c-statistic, calibration slope and calibration intercept. The c-statistic is a non-parametric measure of discrimination, where a score of 1 means the model can perfectly rank individuals according to their predicted risk, whereas 0.5 means discrimination no better than random chance[13]. Model calibration quantifies the alignment between predicted risks (estimated by the model) and the outcome rate observed in the data. A slope of 1 and intercept of 0 describe a perfectly calibrated model[14]. We used Wilcoxon signed-rank tests to calculate median differences in performance between models and between datasets. The negative control (accidental injury) was excluded from these comparisons.

For each outcome, a logistic regression model was fitted in the training set, using the observed binary outcome as the dependent variable and the model's logit-transformed predicted risk as the only predictor variable. Let $\mathrm{s_k}$ denote the logit-transformed predicted risk for outcome $\mathrm{k}$ for a given model, and let $\mathrm{y_k}$ denote the log-odds of outcome $\mathrm{k}$. The recalibration model takes the form:

$$yk = \alpha_k + \beta_k \cdot s_k, k = 1, \dots, 23$$

This produces an outcome-specific recalibration intercept ($\alpha_k$) and slope ($\beta_k$). For each participant in the test set, the recalibrated predicted risk was then calculated as:

$$s_{k,recal} = \alpha_k + \beta_k \cdot s_k$$

The predicted risk ($\mathrm{p_k}$) is then calculated using the inverse of the logit transformation

$$p_k = \frac{1}{1 + e^{-s_{k,recal}}}$$

This was repeated separately for each model (QRISK3, PCE, SCORE2 and SCORE2-OP). As this transformation is monotonically increasing, the rank order of predicted risks is preserved, which leaves the model discrimination (c-statistic) unchanged. Model calibration (slope and intercept) was assessed in the test set before and after recalibration. 95% confidence intervals for the calibration intercept and calibration slope were calculated using the Wald method.

In the permuted feature importance analysis, we first estimated the baseline c-statistic for each outcome on the original test set. For each feature, we randomly shuffled its values for all participants in the test-set (independently of the other variables) and recalculated the c-statistic on the permuted data. To achieve stable results, we repeated this cycle 50 times per feature. We defined the decrease in mean c-statistic after a variable was shuffled as the permuted feature importance for that variable. We set a random seed to ensure the findings were reproducible.

**Sensitivity and Subgroup analysis:**

*Model performance over follow up time:*

We aimed to investigate whether model discrimination changed with the length of follow-up used. To investigate we ran a linear regression with the follow up time as the independent variable and the c-statistic as the dependent variable. The equation for this linear model equation is given by:

$$C_{k,t} = \gamma_k + \delta_k \cdot t + \varepsilon_{k,t}$$

where $C_{k,t}$ is the c-statistic for outcome $k$ at follow-up time $t$, $\gamma_k$ is the intercept, $\delta_k$ is the slope quantifying the change in c-statistic per unit increase in follow-up time, and $\varepsilon_{k,t}$ is the error term. A change in c-statistic of 0.02 over follow-up time points was considered to reflect a meaningful difference in discrimination.

*APOE4 carriership definition:*

The *APOE* genotype was derived using two single nucleotide polymorphisms (SNPs), rs429358 and rs7412, from UKB's whole exome sequencing (WES) data. These two SNPs were used to define carriership of $\varepsilon 2$, $\varepsilon 3$, and $\varepsilon 4$ alleles. Participants were classified as carriers of *APOE4* if they possessed at least one ε4 allele (genotypes ε2ε4, ε3ε4, or ε4ε4) and as non-carriers otherwise. Homozygous carriers of *APOE4* were defined as having two ε4 alleles and heterozygous carriers were defined as having only one ε4 allele. As the ε1 allele is very rare, ambiguous genotypes (ε1ε4 / ε1ε3) were assigned to ε2ε4. Data were extracted for all UKB participants with available WES, regardless of quality control status or ancestry. Participant counts by *APOE4* carrier status are shown in eTable13.

*Study registration:*

This study was not registered nor was a study protocol developed.

*Software:*

All figures in the supplementary material were created using the plot-misc library[15] using Python version 3.1.1. and ggplot2[16] using R version 4.3.1.

**Supplementary Figures**

Figure S1: C-statistics of the repurposed CVD models for neurocognitive outcomes by *APOE4* carrier status.

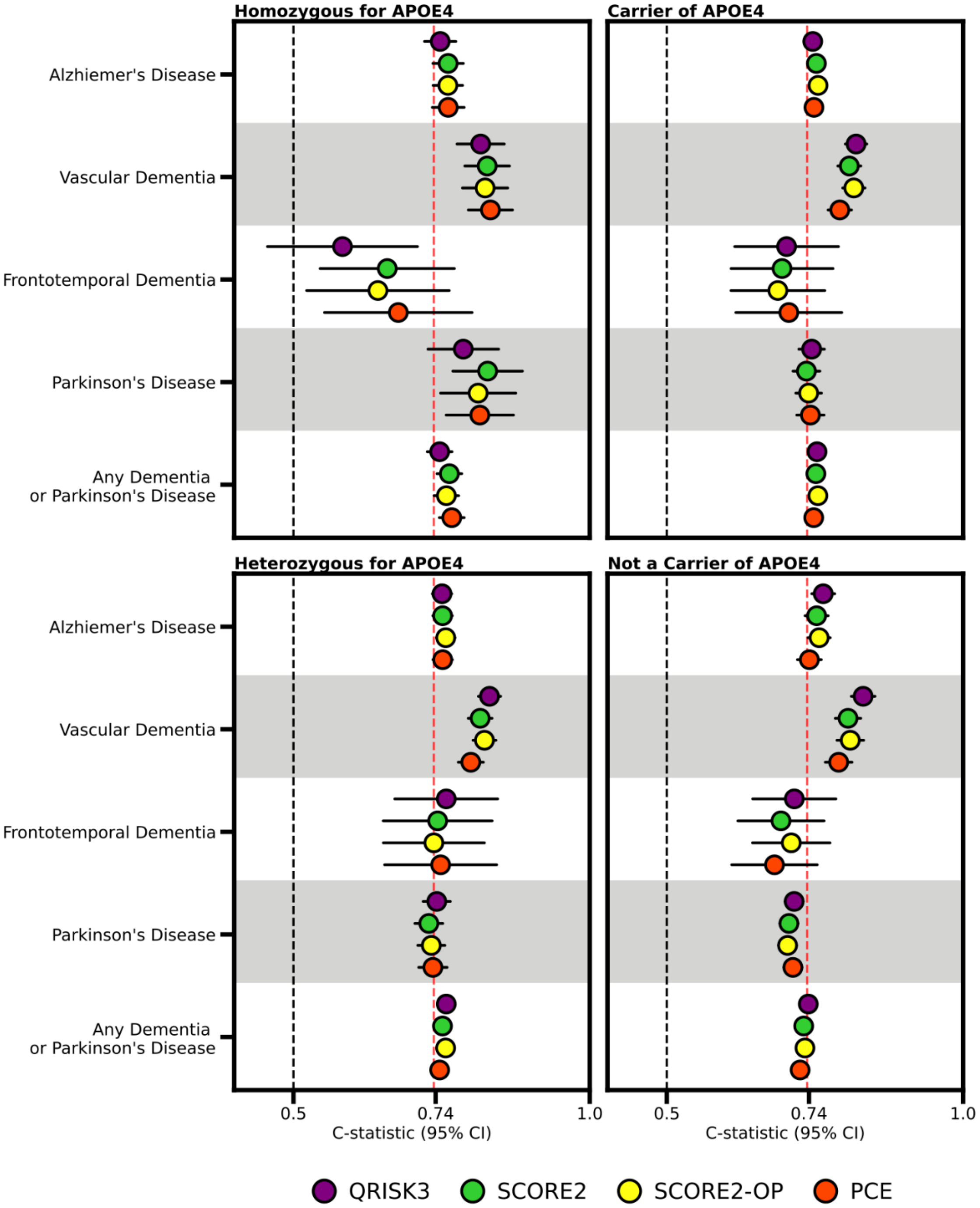

*Forest plot of c-statistics for the four repurposed CVD models for the neurocognitive outcomes in the UK Biobank cohort. The red, vertical dashed line is included as a reference to indicate the performance of QRISK3 for CVD at 10 years (c-statistic of 0·74, 95%CI 0.73; 0.74). Depending on the outcome being predicted, people with pre-existing dementia, Parkinson's disease or both were excluded from the analysis; see eTable12. Abbreviation: 95% CI, 95% confidence interval.*

Figure S2: Feature importance for the Pooled Cohort Equations model.

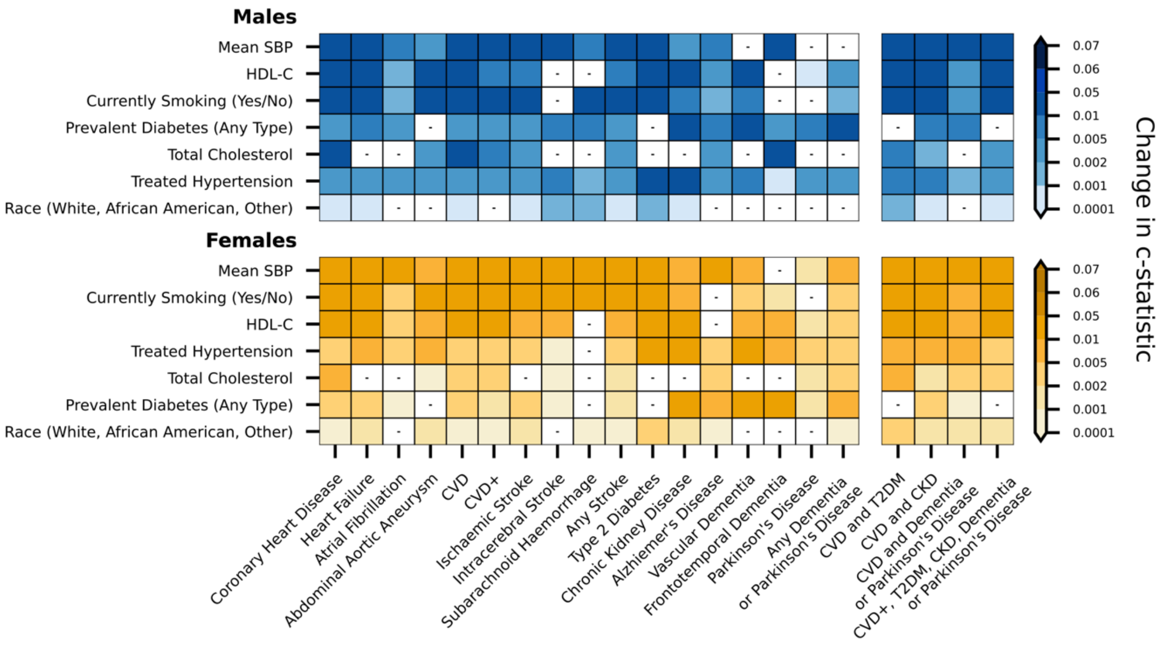

*The feature importance results for the Pooled Cohort Equations model in the UK Biobank for each outcome considered in the study. The top panel (blue) shows results for male participants, and the bottom panel (yellow) shows results for female participants. The predictor variables (y-axis) are ordered by overall importance across all outcomes. The darker shading corresponds to a greater change in c-statistic when that feature was randomly permuted, indicating a greater feature importance. The white tiles with a small, black horizontal line indicate which features did not contribute to the c-statistic in the PCE model for that outcome. Abbreviations: AF, atrial fibrillation; SBP, systolic blood pressure; CKD, chronic kidney disease; CVD, cardiovascular disease; HDL-C, high-density lipoprotein cholesterol; T2DM, type 2 diabetes mellitus.*

Figure S3 Permuted feature importance for the SCORE2 model.

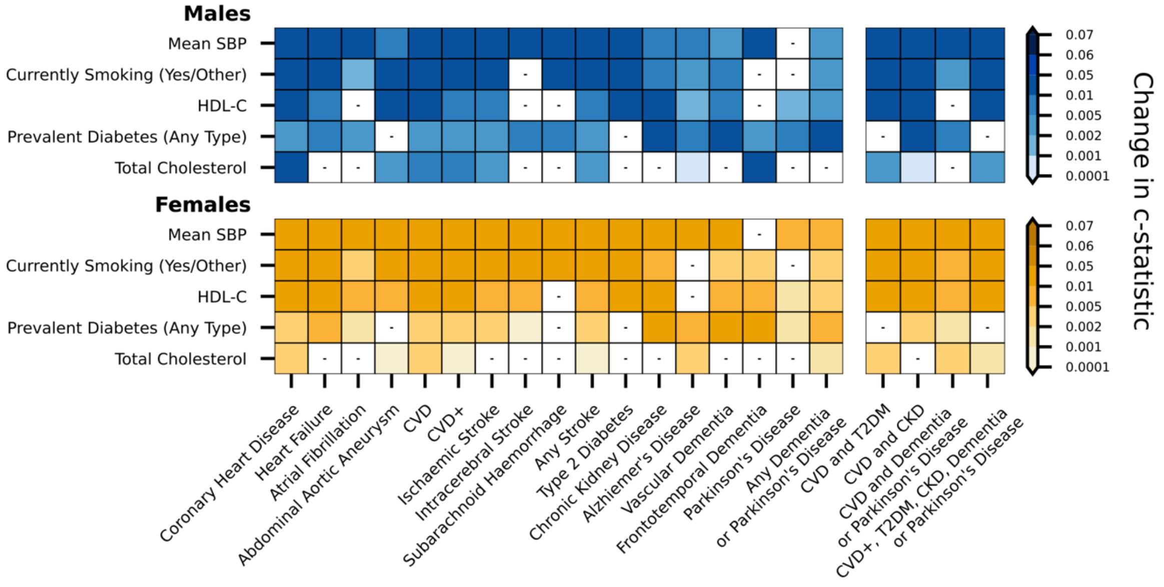

*The feature importance results for the SCORE2 model in the UK Biobank for each outcome. The top panel (blue) shows results for male participants, and the bottom panel (yellow) shows results for female participants. The predictor variables (y-axis) are ordered by overall importance across all outcomes. The darker shading corresponds to a greater change in c-statistic when that feature was randomly permuted, indicating a greater feature importance. The white tiles with a small, black horizontal line indicate which features did not contribute to the c-statistic in the SCORE2 model for that outcome. Abbreviations: AF, atrial fibrillation; CKD, chronic kidney disease; CVD, cardiovascular disease; HDL-C, high-density lipoprotein cholesterol; T2DM, type 2 diabetes mellitus.*

Figure S4: Permuted feature importance for the SCORE2-OP model.

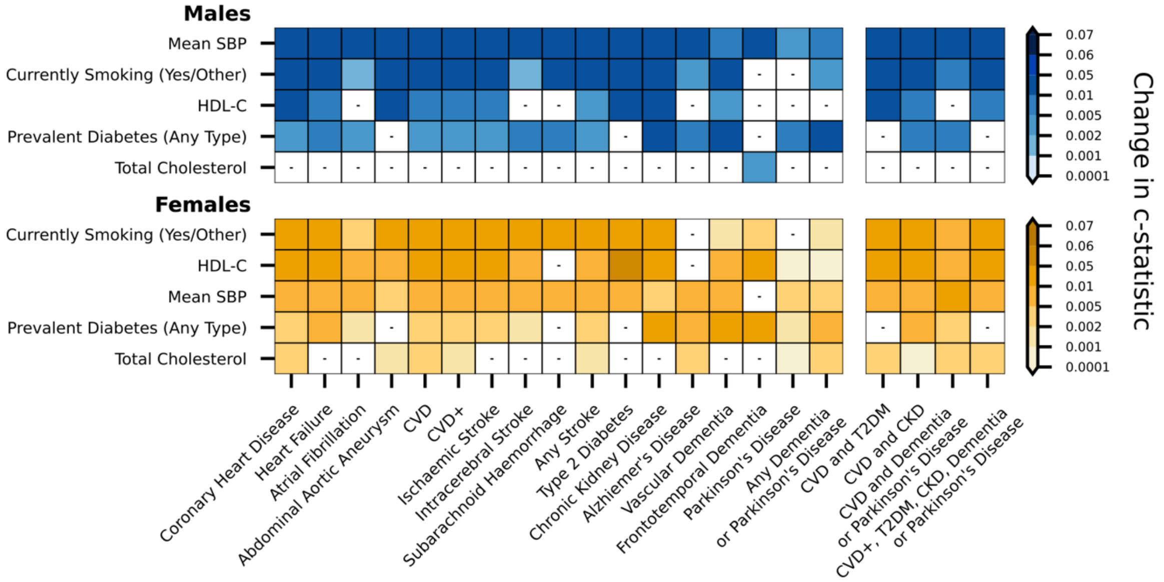

*The feature importance results for the SCORE2-OP model in the UK Biobank for each outcome. The top panel (blue) shows results for male participants, and the bottom panel (yellow) shows results for female participants. The predictor variables (y-axis) are ordered by overall importance across all outcomes. The darker shading corresponds to a greater change in c-statistic when that feature was randomly permuted, indicating a greater feature importance. The white tiles with a small, black horizontal line indicate which features did not contribute to the c-statistic in the SCORE2-OP model for that outcome. Abbreviations: AF, atrial fibrillation; CKD, chronic kidney disease; CVD, cardiovascular disease; HDL-C, high-density lipoprotein cholesterol; T2DM, type 2 diabetes mellitus.*

Figure S5: Calibration plots comparing the repurposed CVD risk models for heart failure, atrial fibrillation, abdominal aortic aneurysm, peripheral arterial disease, and CVD in UK Biobank data after model recalibration.

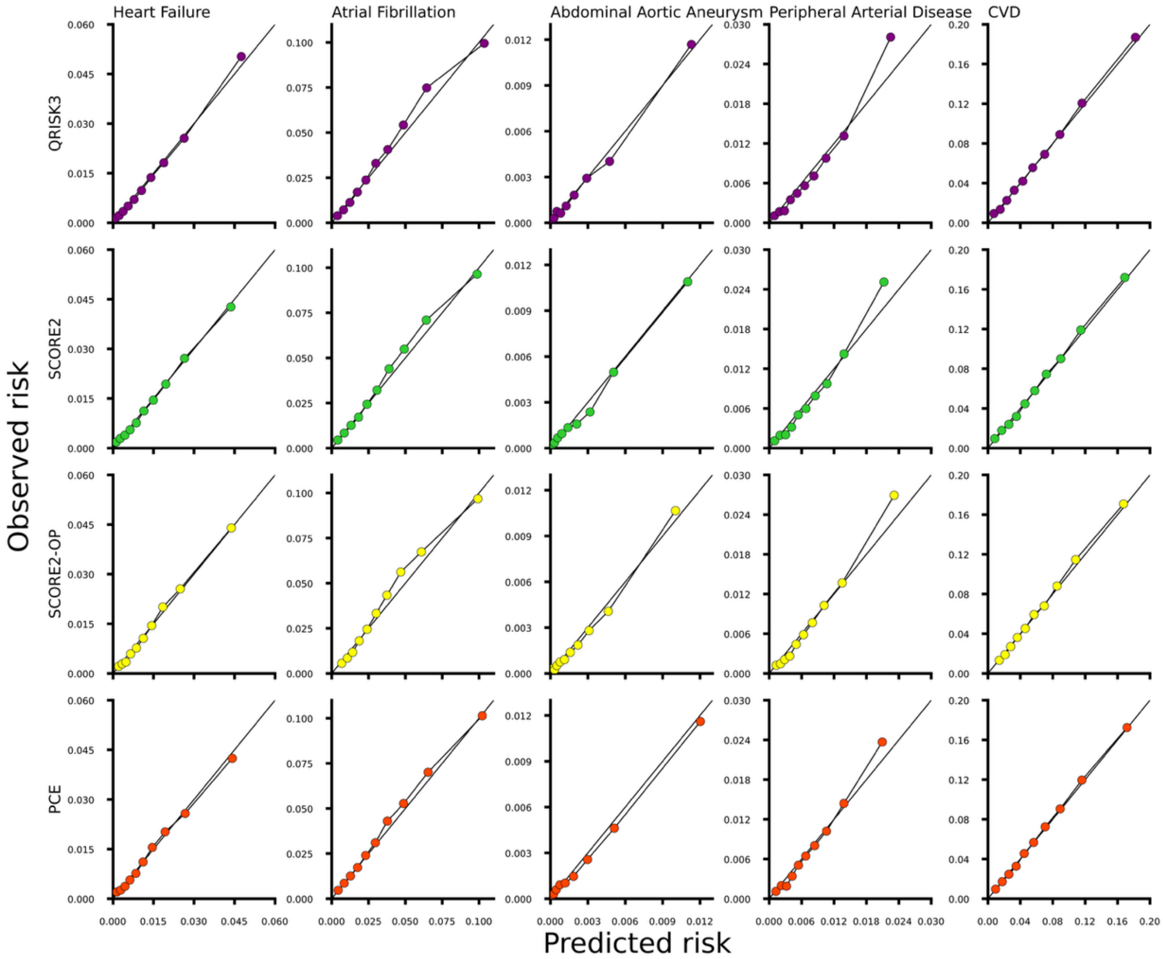

*Agreement between the predicted (x-axis) and observed risks (y-axis) of four CVD risk models (QRISK3, PCE, SCORE2, SCORE2-OP) for heart failure, atrial fibrillation, abdominal aortic aneurysm, peripheral arterial disease, and CVD. Each point corresponds to a risk decile and the solid diagonal line to perfect calibration. For each outcome, participants with prevalent disease were excluded; eTables12. Abbreviations: CVD, cardiovascular disease,*

Figure S6: Calibration plots comparing the repurposed CVD risk models for coronary heart disease, ischaemic stroke, intracerebral stroke, subarachnoid haemorrhage, and any type of stroke in UK Biobank data after model recalibration.

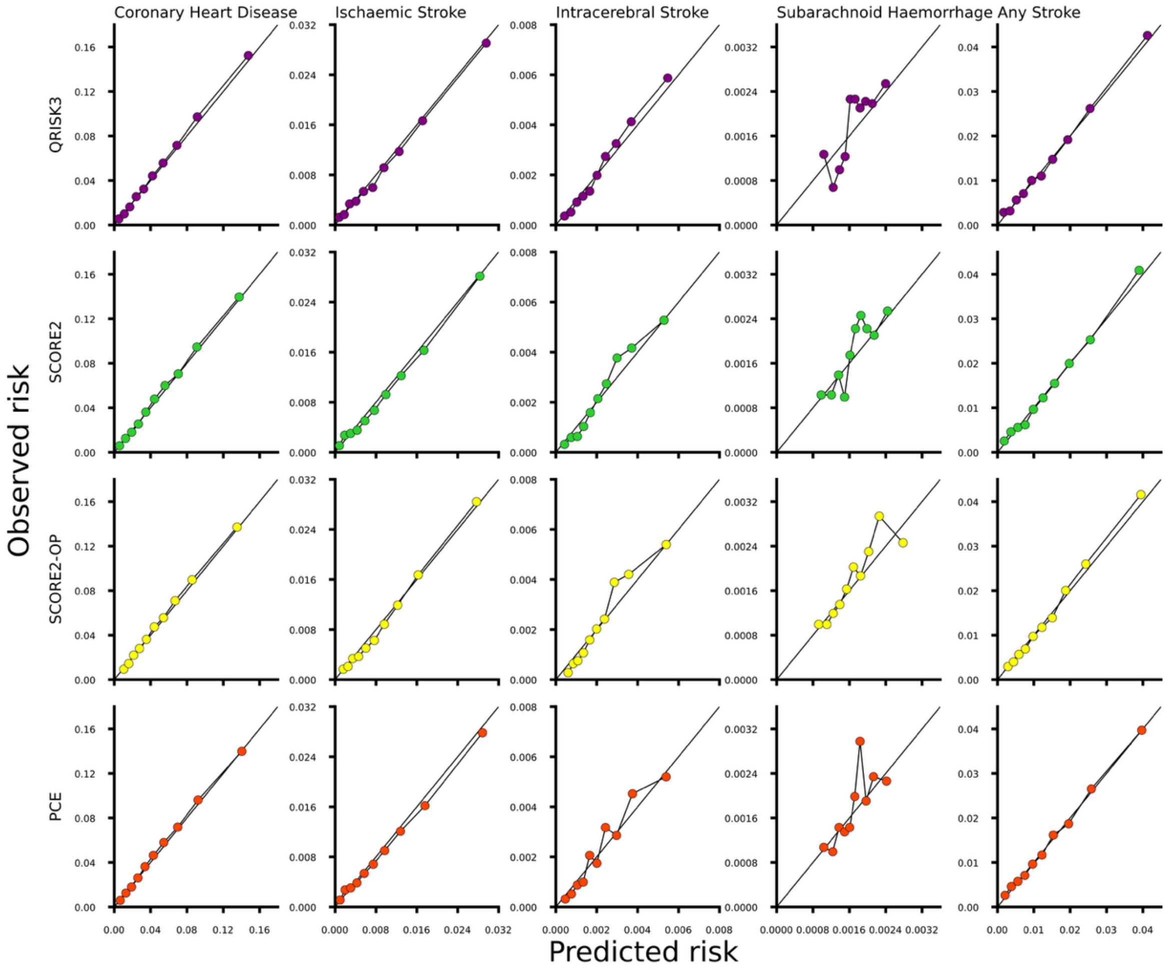

*Agreement between the predicted (x-axis) and observed risks (y-axis) of four CVD risk models (QRISK3, PCE, SCORE2, SCORE2-OP) for coronary heart disease, ischaemic stroke, intracerebral stroke, subarachnoid haemorrhage, and any stroke. Each point corresponds to a risk decile and the solid diagonal line to perfect calibration. For each outcome, participants with prevalent disease were excluded; eTable12.*

Figure S7: Calibration plots comparing the repurposed CVD risk models for Alzheimer's disease, Parkinson's disease, vascular dementia, frontotemporal dementia, any dementia or Parkinson's disease in UK Biobank data after model recalibration.

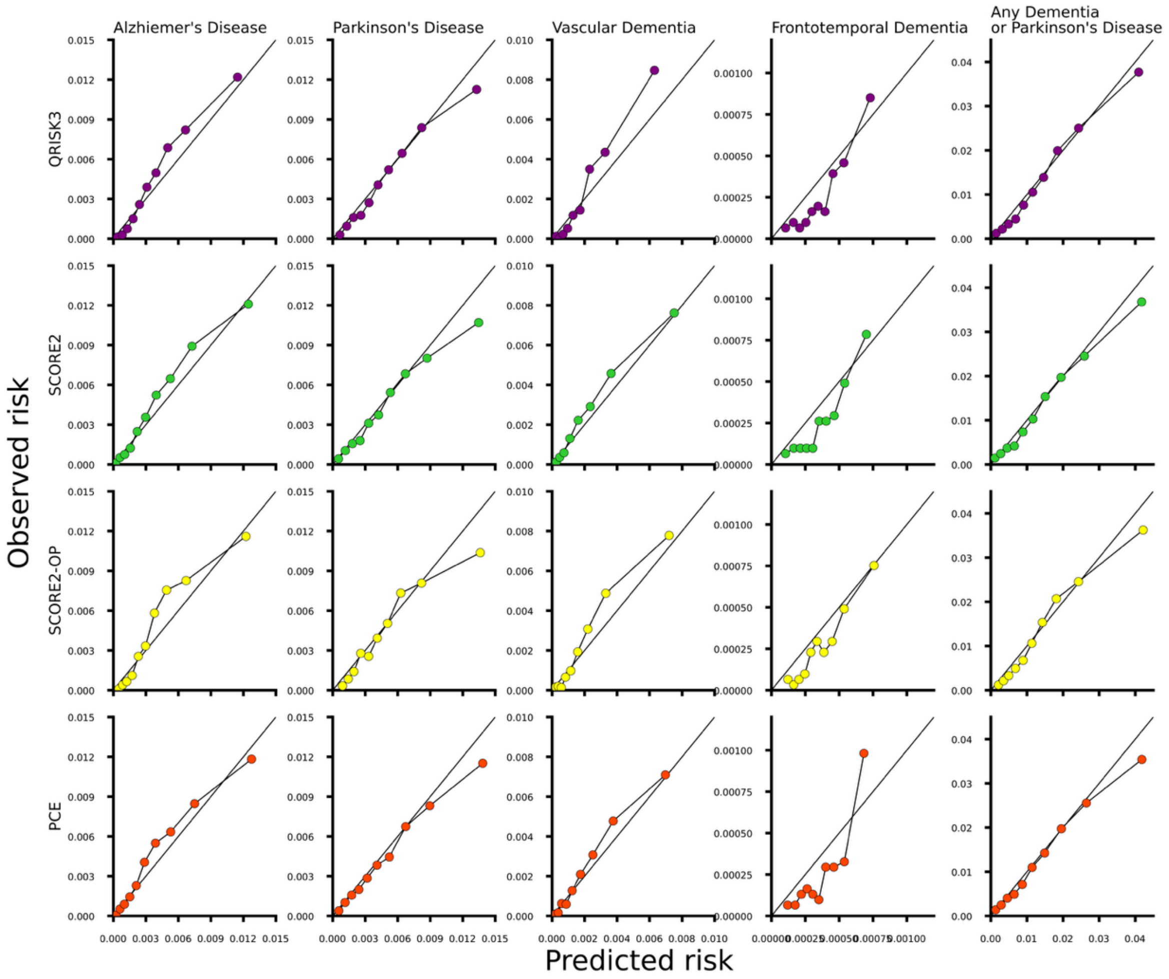

*Agreement between the predicted (x-axis) and observed risks (y-axis) of four CVD risk models (QRISK3, PCE, SCORE2, SCORE2-OP) for Alzheimer's disease, Parkinson's disease, vascular dementia, frontotemporal dementia, any dementia or Parkinson's disease. Each point corresponds to a risk decile and the solid diagonal line to perfect calibration. For each outcome, participants with prevalent disease were excluded; eTable12.*

Figure S8: Calibration plots comparing the repurposed CVD risk models for the CVD-multimorbidity and composite endpoints in UK Biobank data after model recalibration.

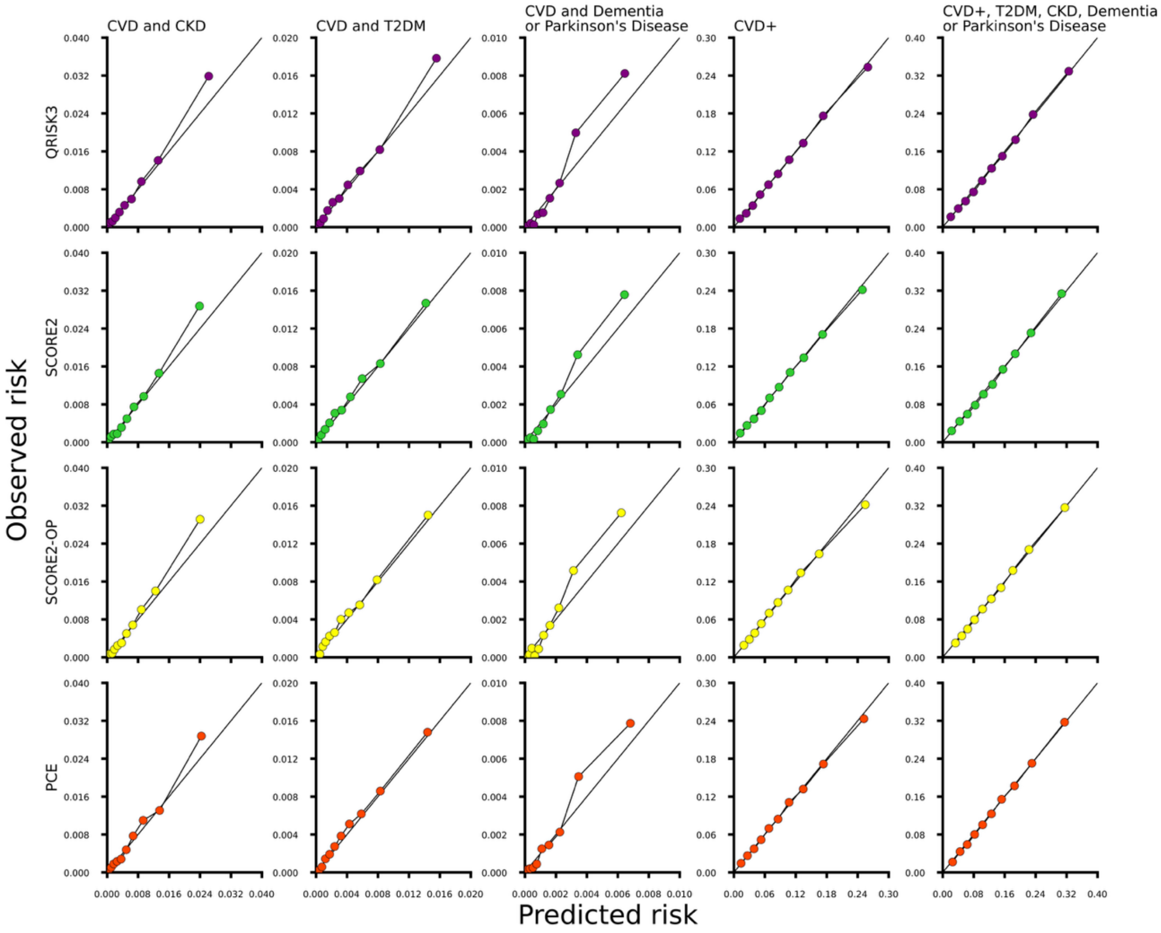

*Agreement between the predicted (x-axis) and observed risks (y-axis) of four CVD risk models (QRISK3, PCE, SCORE2, SCORE2-OP) for the multimorbidity endpoints (CVD and CKD; CVD and T2DM; CVD and any dementia or Parkinson's disease) and the composite endpoints (CVD+ and MACNO). Each point corresponds to a risk decile and the solid diagonal line to perfect calibration. For each outcome, participants with prevalent disease were excluded; eTable12. Abbreviations: Chronic kidney disease, CVD, cardiovascular disease, MACNO; major adverse cardiometabolic or neurocognitive outcome; T2DM, type 2 diabetes mellitus.*

Figure S9: Calibration plots comparing the repurposed CVD risk models for the individual cardiometabolic disease outcomes in UK Biobank data after model recalibration.

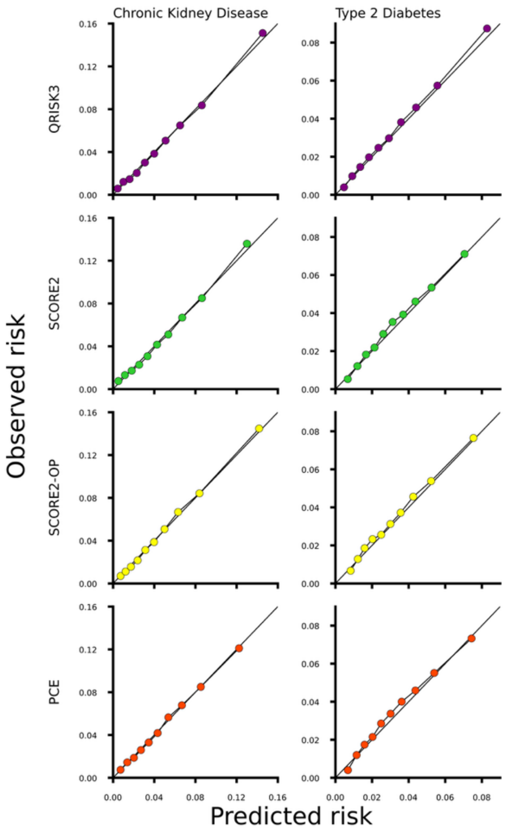

*Agreement between the predicted (x-axis) and observed risks (y-axis) of four CVD risk models (QRISK3, PCE, SCORE2, SCORE2-OP) for T2DM and CKD in the UK Biobank after recalibration. Each point corresponds to a risk decile and the solid diagonal line to perfect calibration. For each outcome, participants with prevalent disease were excluded; eTable12. Abbreviations: Chronic kidney disease, CVD, T2DM, type 2 diabetes mellitus.*

Figure S10: Flow chart showing the counts of included participants at each stage of processing the data to predict incident chronic kidney disease in the UKB.

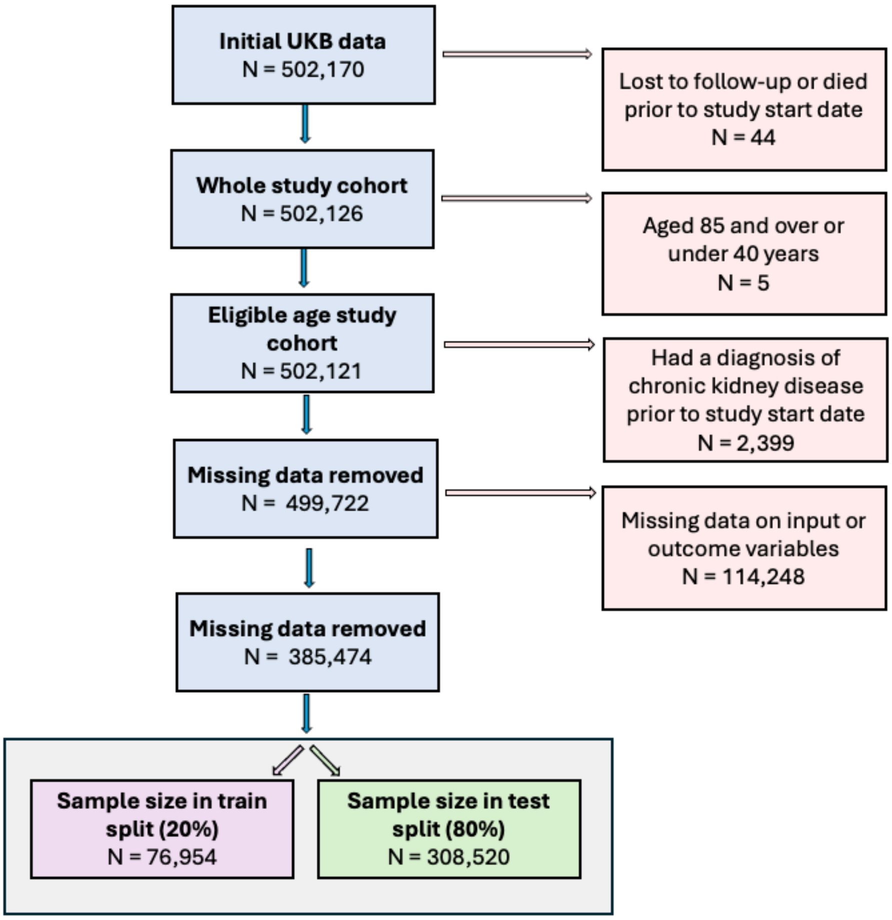

*Flow chart summarising the number of participants excluded while preparing the UK biobank cohort to investigate model performance on predicting incident chronic kidney disease.*

Figure S11: Flow chart showing the counts of included participants at each stage of processing the data to predict incident chronic kidney disease in the original and imputed CPRD cohorts.

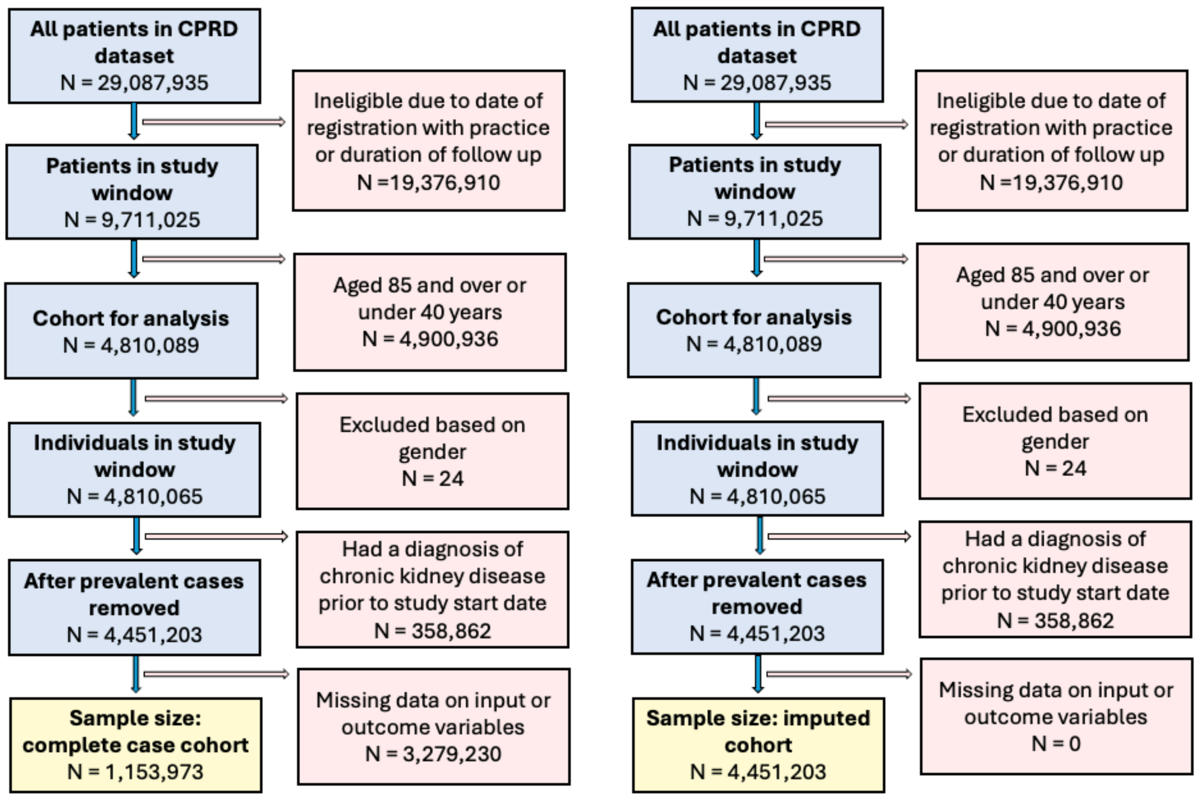

*Flow chart summarising the number of participants excluded while preparing the original (left and imputed (right) CPRD cohorts to investigate model performance on predicting incident chronic kidney disease.*
